# Complementary and alternative medicine use among patients with tuberculosis at a tertiary hospital in Sierra Leone: A cross-sectional study

**DOI:** 10.64898/2026.07.01.26357075

**Authors:** Iye Pateh Jalloh, Solomon F. F. Sandy, Mary S. Kanu, Zaratu Sankoh, Fatmata Batuly Bah, Alieu Kanu, Alhassan Barrie, Fatima Jalloh, Isaac O. Smalle, Onome T. Abiri, Sulaiman Lakoh, Mohamed B. Jalloh, Mamadu Baldeh, Alhaji U. Njai, Abdul Karim Bah

**Affiliations:** Connaught Hospital, University of Sierra Leone Teaching Hospitals Complex, Freetown, Sierra Leone; Institute for Global Health, Faculty of Population Health Sciences, University College London, London, United Kingdom; Alliance for Medical Research in Africa (AMedRA), Dakar, Senegal; Medical Research Council Gambia at London School of Hygiene and Tropical Medicine, London, United Kingdom; Koinadugu College, Kabala, Sierra Leone; California University of Science and Medicine, Colton, California, United States of America; College of Medicine and Allied Health Sciences, University of Sierra Leone, Freetown, Sierra Leone

## Abstract

**Background:** Complementary and alternative medicine (CAM) is widely used alongside biomedical treatment in sub-Saharan Africa, yet evidence among patients with tuberculosis (TB) is limited. Concurrent herbal and anti-TB drug use raises concerns about safety, communication, and symptom attribution. We aimed to estimate the prevalence and describe patterns of CAM use, and to explore sociodemographic associations and disclosure practices among adults receiving TB care.

**Methods:** We conducted a cross-sectional study at Connaught Hospital, Freetown, between February and April 2024. Adults (≥18 years) with microbiologically confirmed TB were consecutively enrolled. Structured interviews assessed CAM use in the past 12 months, modalities, motivations, adverse experiences, and disclosure. Prevalence was estimated using descriptive statistics. Multivariable logistic regression explored sociodemographic associations with CAM use.

**Results:** Among 160 participants (median age 35 years; 68.1% male), 109 (68.1%, 95% CI 60.6–74.8) reported CAM use. Herbal remedies were the most common modality (74.3% of users), particularly ginger (34.9%) and garlic (20.2%). The primary motivation was symptom relief (73.4%). Only 27.6% of CAM users disclosed use to a healthcare provider, most commonly because they reported not being asked (57.5%). In multivariable analysis, only student status was independently associated with lower odds of CAM use (aOR 0.16, 95% CI 0.05–0.56).

**Conclusions:** CAM use was common among adults receiving TB care and was characterized by frequent herbal use and low disclosure. Routine, non-judgmental inquiry about CAM use may improve communication and patient safety during TB treatment.

## Introduction

Complementary and alternative medicine (CAM) encompasses diverse health practices, products, and systems not generally considered part of conventional medicine [1]. While the National Center for Complementary and Integrative Health (NCCIH) provides a widely-used taxonomy, CAM practices in Sierra Leone exist within a distinct socio-cultural context where traditional medicine often serves as the first point of care rather than a complement to biomedical treatment. Studies from Sierra Leone and the sub-region have documented high CAM prevalence across diverse patient populations, with use patterns shaped by healthcare access, cultural beliefs, and pluralistic health-seeking behaviour [14]. When used alongside biomedical treatment, these approaches are termed complementary; when used as substitutes, they are described as alternative. In practice, patients frequently combine both without informing clinicians, particularly when conventional therapies are prolonged, carry side effects, or require strict adherence [2, 3].

Tuberculosis (TB) remains a leading infectious cause of death worldwide, with the World Health Organization (WHO) African Region bearing a disproportionate burden. In 2022, an estimated 2.5 million people developed TB in Africa, with approximately 424,000 deaths [4, 5]. Sierra Leone ranks among the high-burden countries in West Africa, facing persistent challenges including delayed diagnosis, stigma, health-system barriers, and treatment interruptions [6].

In resource-constrained, high-burden settings, TB care is frequently shaped by pluralistic health-seeking behaviours, where biomedical treatment coexists with traditional, spiritual, and herbal practices [7, 8]. The prolonged duration of TB treatment, coupled with medication toxicity and socioeconomic pressures, may increase the appeal of readily available remedies as adjuncts to relieve symptoms, boost immunity, or accelerate recovery [22].

The relevance of CAM to TB care extends beyond cultural preference. First, CAM use may influence care pathways, including delayed presentation or interruption of standardised therapy [8, 9]. Second, several herbal preparations have been associated with hepatotoxicity or pharmacokinetic interference, complicating interpretation of adverse effects that overlap with first-line anti-TB drug toxicity [10–12]. Third, non-disclosure creates communication blind spots, limiting clinicians’ ability to counsel on safety, identify interactions, and interpret laboratory abnormalities [2, 3, 13]. These issues are particularly salient in TB, where treatment is prolonged and adverse reactions contribute substantially to non-adherence.

Although CAM use has been documented across multiple chronic and infectious diseases, data specific to TB populations in sub-Saharan Africa remain limited and heterogeneous [8, 9, 14]. Moreover, many studies do not characterise modalities, motivations, or disclosure behaviours, limiting translation into clinical practice.

This study therefore aimed to estimate the prevalence and describe patterns CAM use among adults receiving tuberculosis care at Connaught Hospital, and to explore sociodemographic associations and disclosure practices.

## Methods

### Study design and setting

We conducted a cross-sectional study at Connaught Hospital, the largest tertiary teaching hospital in Freetown, Sierra Leone, between 1 February and 30 April 2024. Connaught Hospital serves as a national referral centre for TB management and provides free diagnostic and treatment services under the National TB Control Programme. The hospital serves a catchment population of approximately 1.5 million people in the Western Area and manages an average of 200 new TB cases annually. The study was designed and reported according to the Strengthening the Reporting of Observational Studies in Epidemiology (STROBE) guidelines [15]. Patients and members of the public were not involved in the design, conduct, or reporting of this research.

### Participants and eligibility criteria

We included adult patients (aged ≥18 years) with microbiologically confirmed pulmonary or extrapulmonary TB attending the outpatient TB clinic during the study period. Microbiological confirmation was defined as positive results from sputum smear microscopy for acid-fast bacilli, GeneXpert MTB/RIF assay, or mycobacterial culture. Patients with multidrug-resistant TB were excluded as their treatment trajectories, duration, and healthcare engagement patterns differ substantially from drug-susceptible TB. Critically ill patients were excluded due to inability to provide reliable recall or informed consent at enrolment.

### Sampling and sample size

Consecutive eligible patients were invited. A theoretical prevalence-based calculation suggested a larger sample, but for feasibility, we recruited 160 participants, providing 95% confidence intervals [CI], with an approximate width of ±7.1%, considered acceptable for exploratory prevalence estimation and descriptive pattern analysis.

For regression, applying the events per variable rule (minimum 10-15 events per predictor variable) [16, 17] with 109 CAM users, the study achieved >15 events per variable, exceeding recommended thresholds.

### Data collection

We developed a structured questionnaire adapted from validated instruments used in previous CAM studies [18]. The questionnaire comprised four domains: sociodemographic characteristics (age, sex, education, employment status); clinical characteristics (TB diagnosis duration, treatment status); CAM use patterns (types, frequency, reasons, sources); and communication with healthcare providers. The questionnaire was pre-tested on 10 patients with TB not included in the final sample, with revisions made for clarity and cultural appropriateness.

Trained research assistants conducted face-to-face interviews in private consultation rooms in English or Krio according to patient preference. Questionnaires were administered electronically using password-protected tablets, with data encrypted during transmission and storage.

### Definitions

CAM was defined according to the National Center for Complementary and Integrative Health as health practices, products, and systems not generally considered part of conventional medicine [1]. CAM use was defined as the consumption or application of non-biomedical therapies with explicit therapeutic intent during the current TB illness episode. Participants were asked whether they had used specific substances as remedies or treatments for TB symptoms or to improve their health condition. Routine dietary consumption of items such as ginger, garlic, or lime in meals was not classified as CAM use; only preparations made or consumed with stated therapeutic purpose (e.g., herbal infusions, concoctions, or remedies obtained from traditional healers) were captured. TB diagnosis was classified as bacteriologically confirmed or clinically diagnosed. Patients were further categorised as new or retreatment. Comorbidities including HIV status, diabetes, and hypertension were ascertained through clinical records and participant self-report.

### Statistical analysis

Continuous variables were assessed for normality using the Shapiro-Wilk test and visual inspection of histograms. Normally distributed variables are presented as mean ± standard deviation; non-normally distributed variables as median with interquartile range (IQR). Categorical variables are presented as frequencies and percentages.

Associations between CAM use and sociodemographic characteristics were examined using Pearson’s chi-square test or Fisher’s exact test when expected cell frequencies were below five. Variables with p<0.20 in univariable analysis were considered for multivariable modelling. Multivariable binary logistic regression explored associations between CAM use and sociodemographic variables. Covariates for multivariable analysis were selected a priori based on available data and established associations with CAM use in the literature. No stepwise or automated variable selection procedures were employed.

Age was collapsed from six categories (18-23, 24-29, 30-35, 36-41, 42-47, >47 years) into three groups (18-29, 30-41, ≥42 years). Education was collapsed from four categories (no formal education, primary, secondary, tertiary) into three groups (no formal/primary, secondary, tertiary). Employment status was collapsed from four categories (employed, student, unemployed, retired) into three groups (employed/retired, student, unemployed), combining employed and retired due to small numbers in the retired category (n=7). Reference categories were selected as the largest groups: age 18-29 years, female gender, no formal/primary education, and employed/retired status.

We assessed multicollinearity using variance inflation factors (VIF<5 considered acceptable). Model fit was assessed using Hosmer-Lemeshow testing and AUC.

Results are presented as adjusted odds ratios (aOR) with 95% confidence intervals. Statistical significance was set at p<0.05. All analyses were performed using Python 3.13 with pandas 2.2, statsmodels 0.14, and scipy 1.13.

### Ethical considerations

Institutional approval was granted by the Faculty of Clinical Sciences, College of Medicine and Allied Health Sciences, University of Sierra Leone (Ref: COMAHS/FCS/D/30). Administrative clearance and permission for data collection were approved by the Hospital Care Manager at Connaught Hospital. All participants provided written informed consent. For participants with limited literacy, research assistants read the consent form aloud and obtained witnessed thumbprints. Confidentiality was maintained by fully de-identifying the dataset. When potentially harmful CAM use was identified, participants were counselled on the risks and referred to their treating physician.

## Results

### Baseline Demographic and Clinical Characteristics of Study Participants

Between February and April 2024, approximately 200 patients attending the TB clinic were screened. Forty patients were excluded, including 15 who were unable to provide informed consent, 12 with multidrug-resistant TB, 8 who were critically ill, and 5 who declined participation. The final analytic cohort therefore comprised 160 participants (**Figure 1**). All participants completed the study questionnaire, and there were no missing data for the primary outcome variables.

**Figure 1.**
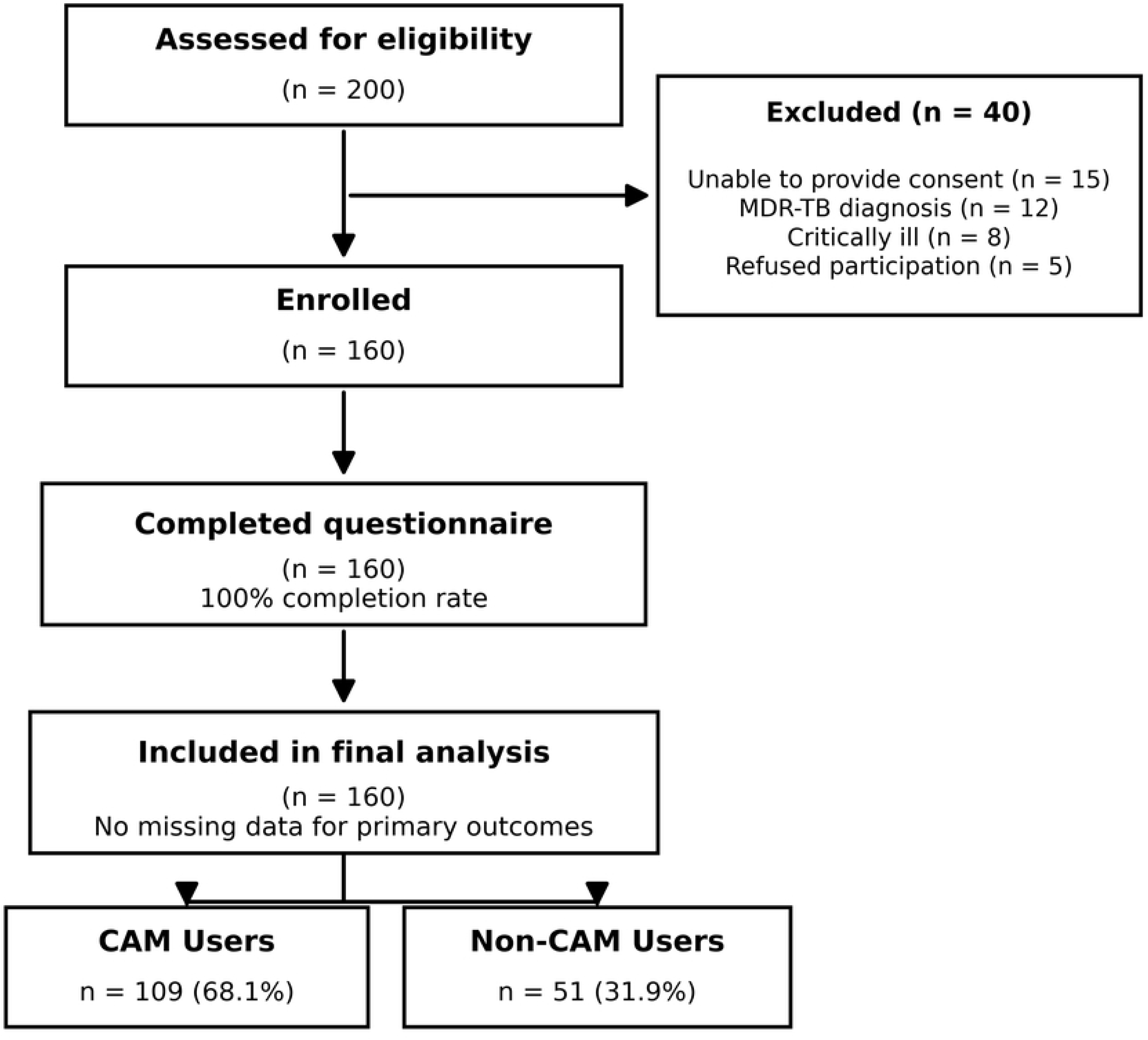
**Study flow diagram showing participant recruitment, exclusions, and final analysis sample according to STROBE guidelines. CAM=complementary and alternative medicine; MDR-TB=multi-drug resistant tuberculosis.**

The median age of the participants was 35 years (interquartile range, 27 to 45). Most participants were male (109 [68.1%]), while 51 (31.9%) were female. With respect to educational attainment, 46 participants (28.7%) had no formal education, 22 (13.8%) had primary education, 70 (43.8%) had secondary education, and 22 (13.8%) had tertiary education. Nearly half of the cohort was employed (74 [46.2%]), whereas 48 (30.0%) were unemployed, 31 (19.4%) were students, and 7 (4.4%) were retired.

Regarding clinical characteristics, 26 participants (16.2%) had been diagnosed with TB within the preceding month, 54 (33.8%) between 1 and 2 months, 41 (25.6%) between 3 and 4 months, 33 (20.6%) between 5 and 6 months, and 6 (3.8%) for more than 6 months. All participants were receiving antituberculosis treatment at the time of enrollment. The baseline demographic and clinical characteristics of the study participants are summarise in **Table 1**.

**Table 1.**
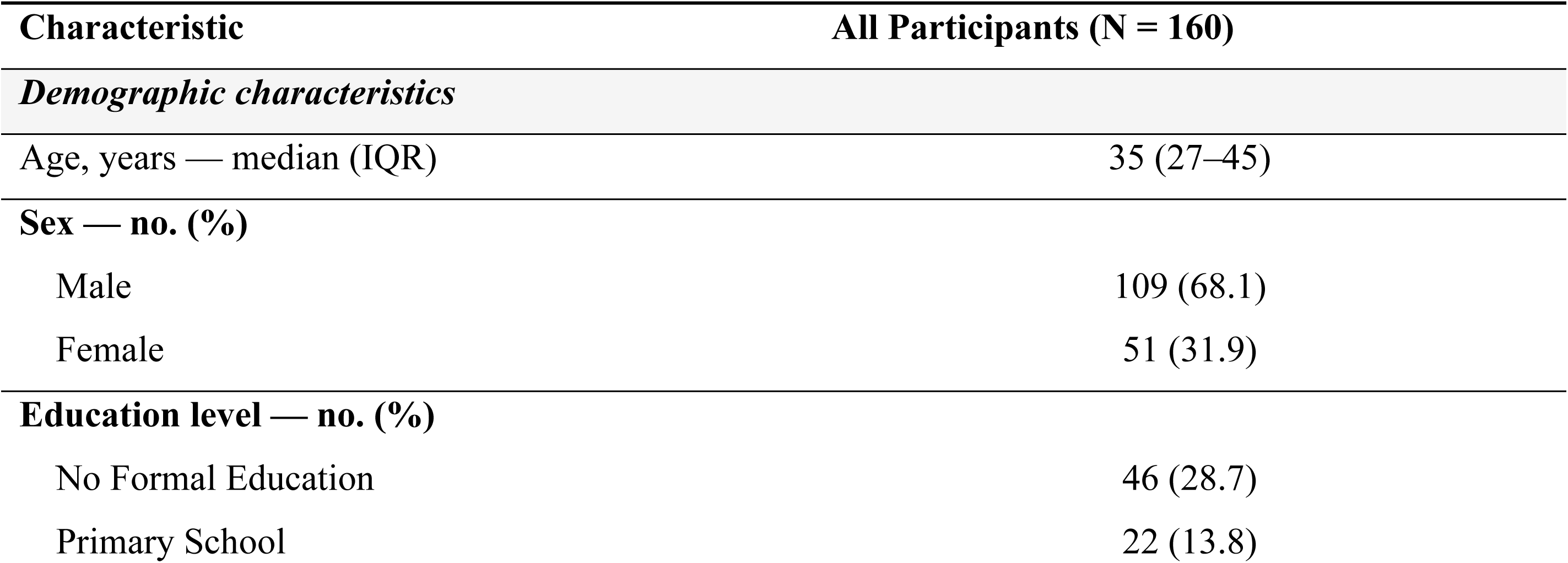
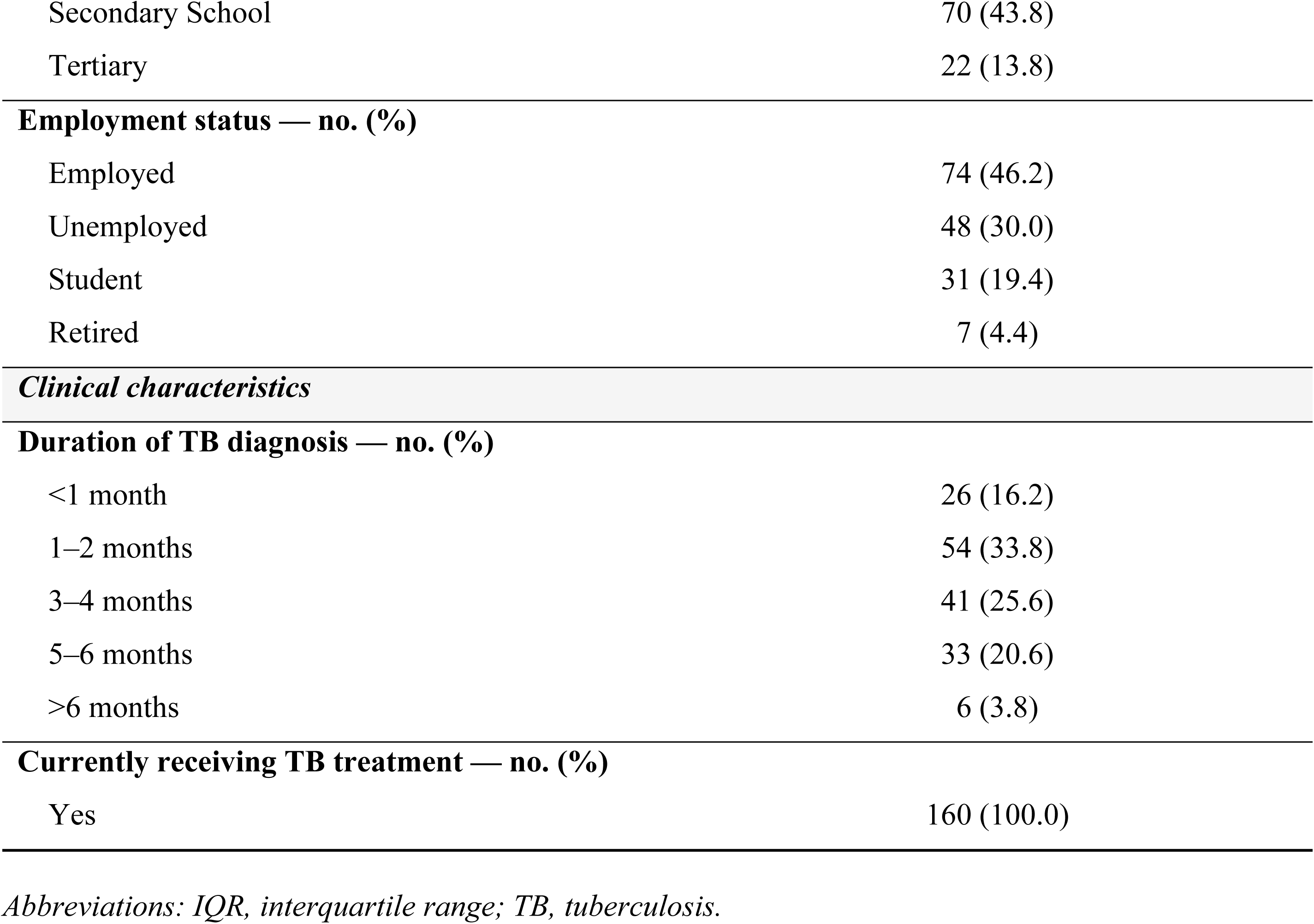
Baseline Demographic and Clinical Characteristics of Study Participants.

### Patterns of Complementary and Alternative Medicine Use

Among the 160 participants, 109 (68.1%) reported CAM use (**Figure 2, Panel A**). The most commonly reported herbal products were ginger (38 [34.9%]), garlic (22 [20.2%]), aloe vera (13 [11.9%]), tea leaf preparations (13 [11.9%]), bitter kola or kolanut (13 [11.9%]), and lime (12 [11.0%]) (**Panel B).** Among CAM users, herbal remedies were the most frequently reported modality, used by 81 participants (74.3%), followed by dietary supplements in 35 (32.1%), prayer or spiritual practices in 26 (23.9%), mind–body practices in 10 (9.2%), and traditional medicine in 1 participant (0.9%) (**Panel C**). The primary reason for CAM use was symptom relief, reported by 80 participants (73.4%). Other reasons included perceived improvement in overall health (32 [29.4%]), immune system support (13 [11.9%]), reduction of medication side effects (12 [11.0%]), and a sense of personal control over health (7 [6.4%]) (**Panel D).**

**Figure 2.**
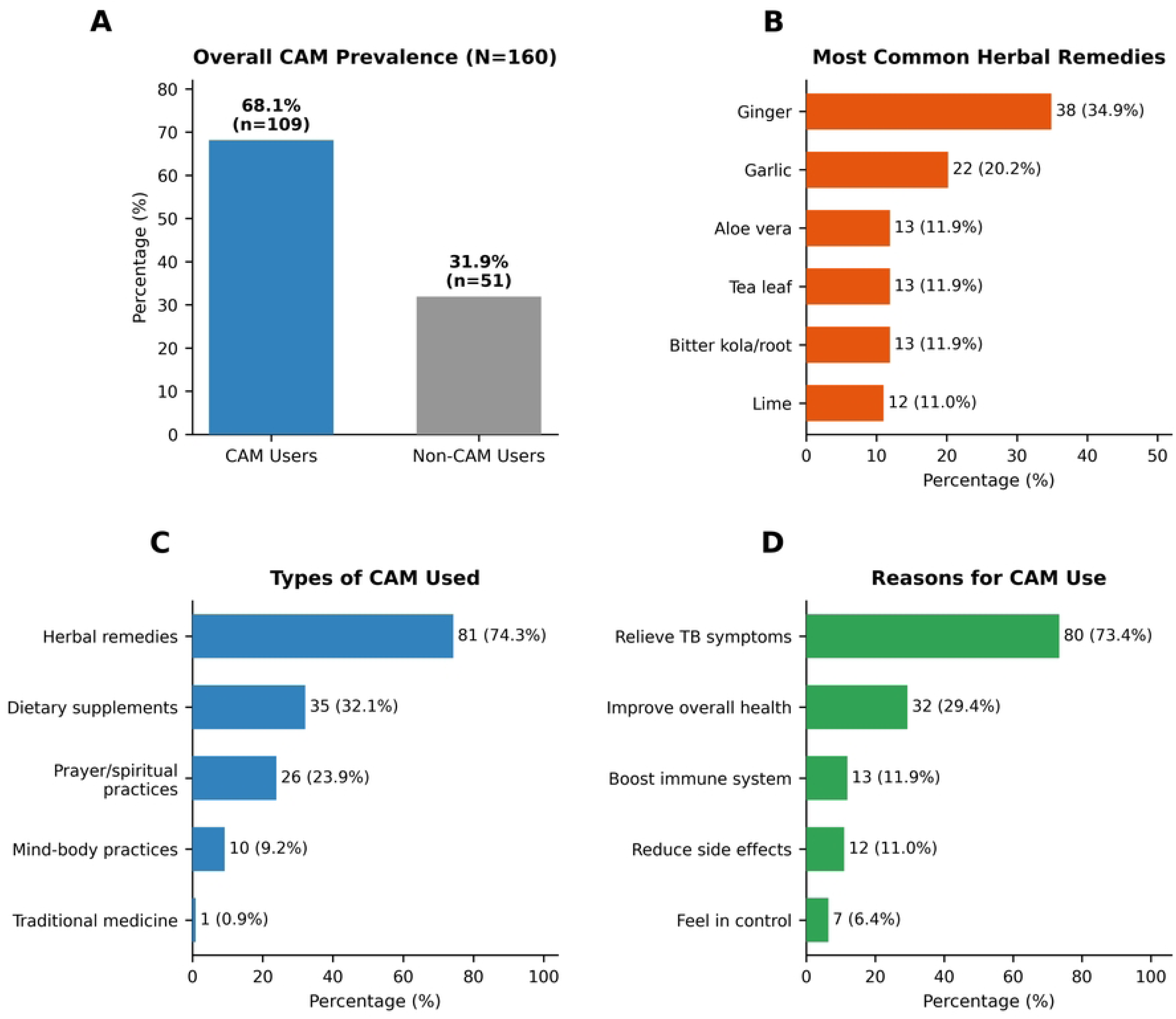
Patterns of Complementary and Alternative Medicine Use among Patients with Tuberculosis. **Panel A** shows the prevalence of CAM use in the study cohort. **Panel B** shows the most commonly reported herbal remedies. Panel C shows the distribution of CAM modalities used among CAM users. **Panel D** shows the reasons for CAM use. Percentages are calculated on the basis of the number of CAM users in each panel, unless otherwise indicated.

### Disclosure to healthcare providers

Among the 109 participants who reported using CAM, only 29 (27.6%) disclosed such use to a health care provider, whereas 80 (72.4%) did not disclose this information (**Figure 3**).

**Figure 3.**
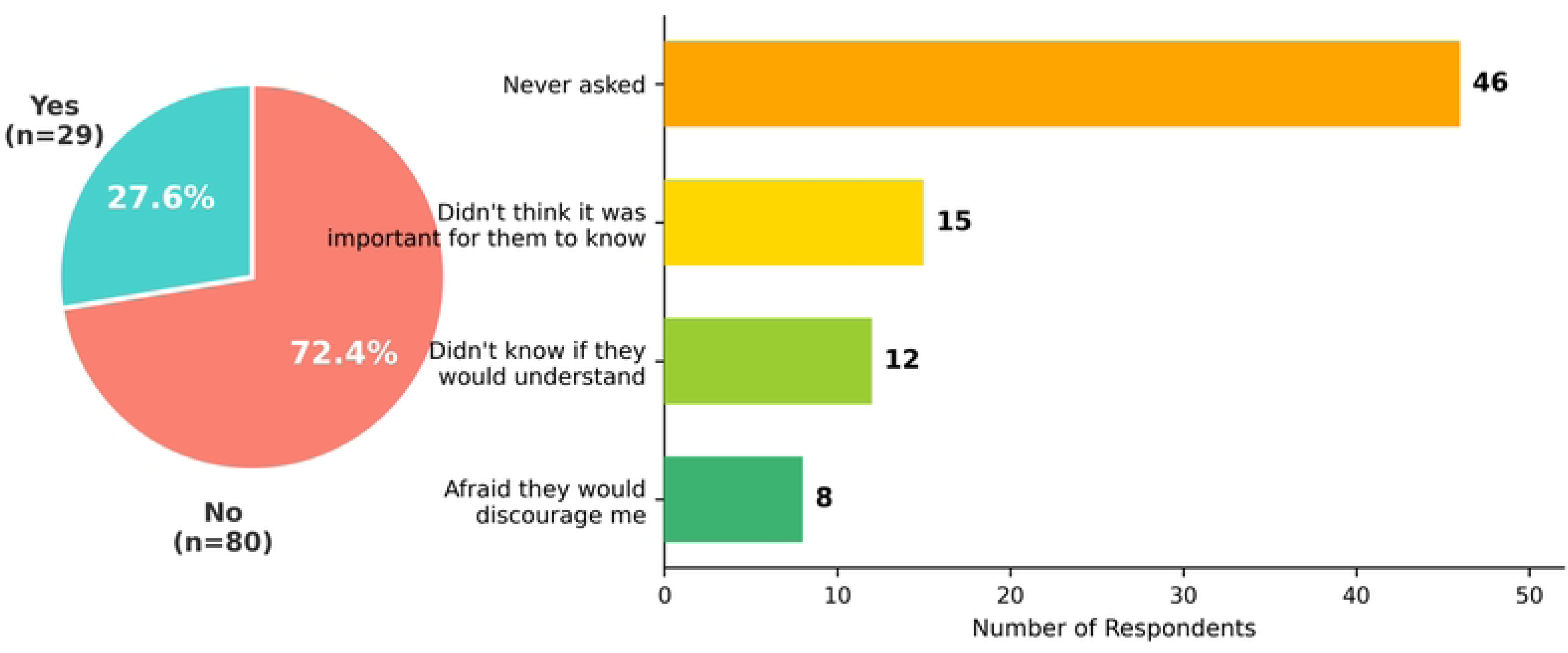
Disclosure of Complementary and Alternative Medicine Use to Health Care Providers. Shown are the proportions of participants who reported using complementary and alternative medicine (CAM) and who did or did not disclose such use to a health care provider, together with the reported reasons for non-disclosure among those who did not inform a provider. Bars indicate numbers of respondents.

Among the 80 participants who did not disclose CAM use, the most frequently cited reason was that they were never asked by a provider (46 [57.5%]). Other reasons included the perception that disclosure was not important (15 [18.8%]), concern that the provider would not understand (12 [15.0%]), and fear of discouragement or a negative response (8 [10.0%]).

### Association between sociodemographic factors and CAM use

In univariate analyses, employment status was significantly associated with the use of CAM (P = 0.013). CAM use was more common among employed participants (57 of 109 [52.3%]) than among non-CAM users (17 of 51 [33.3%]), whereas students were less likely to use CAM (14 of 109 [12.8%] vs. 17 of 51 [33.3%]). No significant differences in CAM use were observed according to age group (P = 0.40), sex (P = 1.00), or educational attainment (P = 0.24).

Among age groups, CAM use was reported by 37 of 109 participants (33.9%) aged 18 to 29 years, 31 (28.4%) aged 30 to 41 years, and 41 (37.6%) aged 42 years or older, with no significant between-group differences. The distribution of CAM use did not differ by sex, with similar proportions among women (35 of 109 [32.1%]) and men (74 of 109 [67.9%]) (P = 1.00). Educational level was also not significantly associated with CAM use, with the highest proportion among those with secondary education (50 of 109 [45.9%]) **(Table 2).**

**Table 2.**
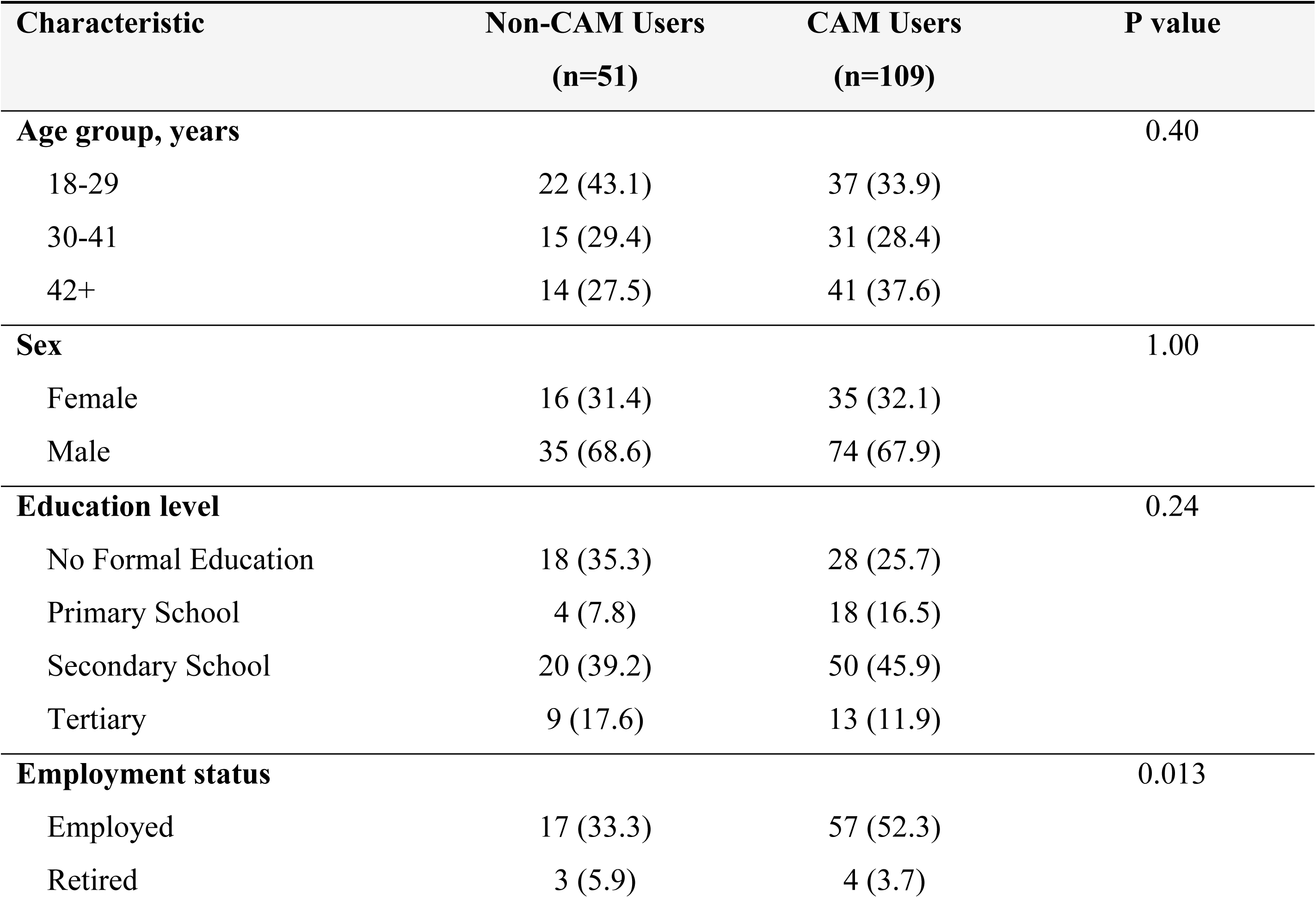
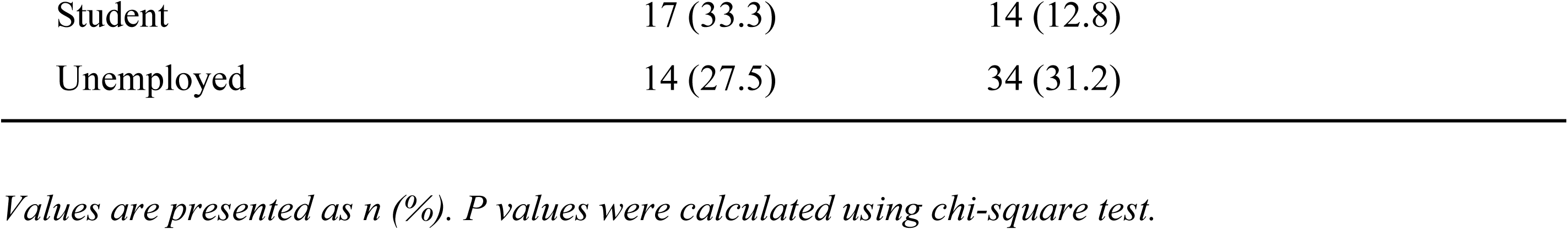
Univariate Analysis of Factors Associated with CAM Use.

In multivariable analysis, only student status was associated with lower CAM use (aOR, 0.16; 95% CI, 0.05 to 0.56; P = 0.004), indicating substantially lower odds of CAM use compared with employed participants (Figure 4).

**Figure 4.**
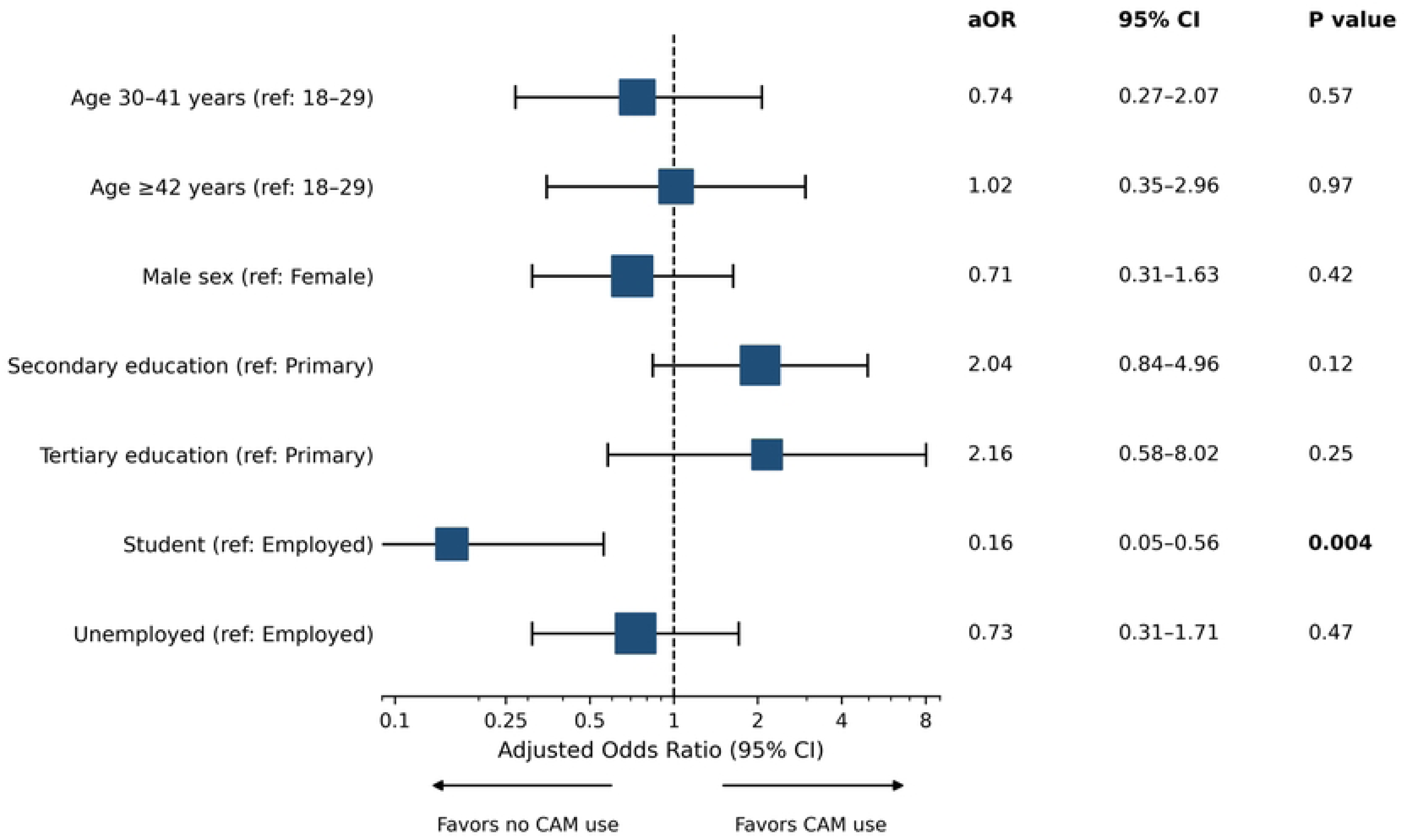
Multivariable Associations between Sociodemographic Characteristics and Use of Complementary and Alternative Medicine. Forest plot showing adjusted odds ratios (aOR) with 95% confidence intervals from multivariable logistic regression analysis of factors associated with the use of complementary and alternative medicine (CAM) among patients with tuberculosis (n=160). Reference categories: age 18–29 years, female gender, no formal/primary education, employed/retired status. The vertical dashed line represents null effect (aOR=1·0). Only student status was significantly associated with CAM use (p<0·05). Model fit: Hosmer-Lemeshow χ²=9·59, p=0·295; AUC=0·63.

Other sociodemographic variables were not significant. Model discrimination was modest (AUC 0.63), indicating limited explanatory power.

## Discussion

This study shows that CAM use is common among adults receiving TB care at Connaught Hospital in Freetown, Sierra Leone, dominated by herbal remedies and characterised by low disclosure, largely because patients reported that clinicians did not ask. The observed prevalence exceeds many regional estimates and may reflect pluralistic health-seeking behaviour. A systematic review of traditional and complementary medicine use in the region reported wide variation across countries and populations but consistently identified herbal medicines as the most commonly used modality [14]. TB care may amplify CAM demand because symptoms can be prolonged, treatment is lengthy, and stigma and uncertainty shape health-seeking behaviours[7, 8]. In pluralistic health systems, patients may move between biomedical and traditional providers without viewing these choices as mutually exclusive. Our finding that symptom relief and wellbeing were dominant motivations suggests CAM is used as an adjunct to conventional care, often addressing unmet symptomatic or psychosocial needs. Notably, substances with dual dietary-medicinal roles (e.g., ginger, garlic) were only classified as CAM when used with explicit therapeutic intent. Kola nut, despite its widespread cultural use in Sierra Leone, had low prevalence as a therapeutic agent in this cohort and did not drive overall CAM estimates.

Non-disclosure of CAM use represents an important actionable finding. Patients perceived that clinicians did not ask, suggesting a missed opportunity for communication rather than confirmed provider behaviour. This pattern mirrors evidence from diverse settings. Systematic reviews report non-disclosure rates ranging from approximately one-quarter to three-quarters, identifying provider non-inquiry, fear of negative responses, and perception that disclosure is unimportant as major barriers [2, 19]. Meta-analytic work emphasises that disclosure is strongly influenced by patient-provider communication quality and whether clinicians ask directly [19]. These data support a practical inference: routine inquiry about CAM is likely to increase disclosure, creating opportunities to identify risks, align expectations, and strengthen therapeutic relationships.

Safety implications require caution. First-line anti-TB regimens can be hepatotoxic and commonly cause gastrointestinal intolerance; concurrent herbal ingestion can complicate symptom attribution and may contribute to preventable treatment interruptions. Systematic reviews describe plausible pharmacokinetic and pharmacodynamic mechanisms for herb-drug interactions, including effects on cytochrome P450 enzymes and drug transporters, though clinical relevance varies by product and context [10, 11]. Separate reviews highlight that herbal and dietary supplements can cause drug-induced liver injury, with causality assessment challenging due to product heterogeneity [12, 20]. In our cohort, 11% of CAM users reported complications, most commonly symptom worsening or diarrhoea. While self-reported and non-specific, these findings underscore the importance of documenting CAM use when evaluating adverse symptoms during TB therapy.

Our results are relevant to adherence and the broader TB care experience. Although adherence was not directly measured, evidence from West Africa indicates that medicinal plant use may be associated with adherence difficulties during TB treatment [21]. The motivations reported here suggest patients may use CAM to manage persistent symptoms, perceived side effects, or illness-related anxiety. This presents a patient-centred opportunity: TB teams can proactively address common symptoms, optimise supportive care, and provide clear guidance on when to seek review, thereby reducing perceived need for unmonitored adjunct therapies. Importantly, counselling should avoid stigmatising language, since fear of discouragement was a reported reason for non-disclosure and is consistently identified in the literature [2, 19].

The lower use among students may reflect different health beliefs or access to biomedical information. The absence of other independent associations and modest model performance suggest unmeasured social and cultural determinants. Future studies should incorporate variables reflecting explanatory models of illness, trust in healthcare, social networks, household income, and access to traditional healers, and should consider qualitative methods to understand decision-making around CAM use during TB treatment.

These findings suggest practical actions for TB services in Sierra Leone and similar contexts. First, integrate CAM assessment into routine history-taking using standardised prompts that normalise disclosure. Second, strengthen provider communication skills for non-judgemental inquiry and culturally respectful counselling. Third, safety-focused counselling should document CAM type and timing, discourage high-frequency use of unknown preparations during intensive therapy, encourage prompt symptom reporting, and consider targeted liver function monitoring when clinically indicated. Finally, TB programmes could develop patient education materials explicitly discussing concurrent herb and TB medication use, framing this as a safety issue rather than moral judgement.

## Strengths and limitations

This study provides detailed characterisation of CAM use among TB patients in Sierra Leone, including modalities, frequency, motivations, disclosure behaviours, and self-reported adverse experiences. Data completeness was high, and inclusion of disclosure and motivation measures strengthens programmatic relevance. Conducting the study at the national adult TB referral centre increases relevance for tertiary TB care in urban Sierra Leone.

Several limitations should be acknowledged. The cross-sectional design precludes causal inference regarding whether CAM use influences adherence or outcomes. CAM use and adverse events were self-reported, subject to recall and social desirability bias. The 12-month recall window included general health CAM use, potentially overestimating TB-specific use. The single-site tertiary setting may not represent rural populations or patients managed outside tertiary care. The exclusion of MDR-TB and critically ill patients may introduce selection bias if CAM use differs systematically in these groups. Patients experiencing treatment failure or prolonged illness may be more likely to seek alternative therapies.

We did not measure adherence, microbiological outcomes, or laboratory markers, limiting our ability to link CAM use to clinical endpoints.

## Conclusions

CAM use among adults receiving TB care at Connaught Hospital in Freetown, Sierra Leone was common and poorly disclosed. Integrating routine, supportive CAM screening into TB services may strengthen patient safety and therapeutic communication.

## Data Availability

The data underlying this study cannot be shared publicly due to ethical and legal restrictions regarding patient confidentiality and the sensitive nature of clinical tuberculosis data. De-identified data are available to qualified researchers upon reasonable request. Data access requests may be directed to the College of Medicine and Allied Health Sciences Research Ethics Committee at.

